# Assessment of somatosensory and cognitive-motor processing time in retired athletes with a history of repeated head trauma

**DOI:** 10.1101/2022.07.20.22277880

**Authors:** Alan J. Pearce, Doug King, Dawson J Kidgell, Ashlyn K Frazer, Mark Tommerdahl, Catherine M Suter

**Author notes:** Corresponding Author: A/Prof Alan Pearce PhD, College of Science, Health and Engineering, La Trobe University, Kingsbury Avenue, Bundoora, Melbourne, Victoria, AUSTRALIA, Twitter: @alanpearcephd.

## Abstract

Measurement of the adverse outcomes of repeated head trauma in contact sport athletes is often achieved using tests where the comparator is the ‘score’ or the ‘accuracy’ obtained. While it is expected that ex-athletes would perform worse than controls, previous studies have shown inconsistent results. Here we have attempted to address these inconsistencies from a different perspective by quantifying not only accuracy, but also the time of motor responses (response time). We tested age-matched control subjects who have never experienced head trauma (n=20; 41.8 ± 14.4 years), and two cohorts of retired contact sport athletes with a history of head trauma and concussions; one with self-reported concerns (n=36; 45.4 ± 12.6 years), and another with no ongoing concerns (n=19; 43.1 ± 13.5 years). Participants performed cognitive (*Cogstate*) and somatosensory (*Cortical Metrics*) testing and both accuracy and response time were recorded. Transcranial magnetic stimulation (TMS) was undertaken to investigate corticospinal conduction and excitability. Results showed that in both test batteries there was little difference between groups when considering only accuracy scores. By contrast, response times in all but one test revealed that ex-athletes with self-reported concerns were significantly slower compared to no concern ex-athlete or control groups (*p* ranges 0.031 to <0.001). TMS latency showed significantly increased conduction time (p=0.008) in the group with ongoing concerns. These findings suggest that incorporating response times in cognitive and somatosensory testing is more informative than considering accuracy scores alone when assessing cognitive processing ability in retired contact sport athletes with ongoing brain health concerns.

## Introduction

The long-term neurological sequelae of head trauma in retired contact sport athletes is of ongoing global concern and investigation. Research into the cognitive and neuropsychological health of retired athletes has by now been carried out over decades, but studies have not always provided consistent results. In particular, there is disparity with respect to self-reported symptomology or concern, and the results of commonly used objective measurements. This is a problem highlighted by Cunningham et al (2020) in a systematic review of 46 cross-sectional studies of retired athletes with a history of sports related concussion. While almost 80% of studies included ex-athletes with self-reported concerns about their cognitive health, only half to two-thirds of these studies showed any impairment in objective measurement of psychomotor function, executive function, or memory.

Assessments of cognitive function generally rely on a performance ‘outcome’, and these can be binary (such as correct detection of something or not), or they can be continuous or additive (such as the number of errors made during a given test) (Harvey, 2019). However, as illustrated by Cunningham et al (2020), reliance on test outcome measures may not detect subtle impairments, particularly in single testing sessions, that reflect self-reported concerns. This implies that the current objective testing regimes that rely purely on outcome measures are insufficient in measuring subtle cognitive processing abilities (De Boeck & Jeon, 2019; Kyllonen & Zu, 2016).

De Boek and Jeon (2019) argue that while cognitive tests measure overall performance abilities, as determined by the number of correct or incorrect responses, less attention is given to quantifying process abilities, reflected by response time. While it is acceptable to determine performance outcome without knowledge of the processes involved, it is only half of the story. Providing cohorts such as retired contact sports athletes who generally are high functioning but struggle with daily activities, an explanation of not only outcomes, but also the process, allows for increased understanding as well as more informative feedback that may assist in interventional therapies (De Boeck & Jeon, 2019).

Response time has long been a consideration in cognitive ability measurements, but the increasing precision in measurement by use of computerised testing has allowed for response time data collection to understand cognitive ability in healthy populations (Kyllonen & Zu, 2016). Consequently, interest in response time has revived, with a number of models being developed for use in psychology (see reviews by Kyllonen and Zu 2016, De Boeck and Jeon 2019). However, the use of response time data as a reflection of processing ability appears to not be utilized in exploring long-term consequences of repeated head trauma in retired athletes (Ebaid et al., 2017). A small number of studies have previously employed psychomotor reaction time in retired athletes (Cunningham et al., 2020). However, reaction time and response time are two distinct variables with the former describing the speed of detecting the stimulus, while the latter describes a speed-accuracy trade-off for the determination of the *correct* response to a given stimulus, rather than simply responding to a stimulus (De Boeck & Jeon, 2019; Kyllonen & Zu, 2016; A. Tommerdahl et al., 2019).

Our previous work on sensorimotor and neurophysiology of individuals with persistent post-concussion symptoms (Pearce et al., 2020), and chronic long-term outcomes in retired athletes (Pearce et al., 2021), demonstrated slower sensorimotor reaction time in symptomatic individuals when compared with controls. Here we present our studies of *response* time in two groups of retired contact sport athletes’: one with ongoing concerns about their cognitive health and the other with no ongoing concerns. We compared these two groups to age-matched controls using two different computerised testing applications. We present both performance outcome and response time data, as well as single pulse transcranial magnetic stimulation (TMS) for quantification of corticospinal excitability.

## Methods

As part of a larger research project, studies reported here were conducted on a convenience sample of 75 male participants (retired contact sport athletes *n*=55; age-matched male controls *n*=20; Table 1). Participants were pre-screened for TMS suitability (Rossi et al., 2011) and provided written informed consent to participate in the study as approved by the University Ethics Committee (HEC18005).

**Table 1.** Participant demographics (mean ± SD)

|  | Age in<br>years | Education<br>(years) | Competitive<br>career (years)* | Number of<br>concussions | Years since last<br>concussion |
| --- | --- | --- | --- | --- | --- |
| **Control**<br>**(n=19)** | 41.8 ±<br>14.4 | 15.6 ± 2.4 | nil | n/a | n/a |
| **No concern**<br>**(n=20)** | 43.1 ±<br>13.5 | 14.8 ± 1.9 | 24.6 ± 8.3 | 5.2 ± 4.1 | 13.3 ± 7.8 |
| **Self-concern**<br>**(n=36)** | 45.4 ±<br>12.6 | 14.3 ± 2.7 | 25.1 ± 9.6 | 6.5 ± 4.1 | 16.5 ± 10.7 |

\* Includes competitive junior career

The retired playing group were divided into sub-groups based on their self-reported *fatigue and related symptoms* score (Johansson et al, 2009, see section ‘symptom self-report’): participants with ongoing self-reported concerns regarding their mental and cognitive health relating to their history of head trauma experienced in sport (‘self-concern’: n=36), and those who acknowledged they had a history of head trauma from sport but did not express any enduring concerns (‘no concern’: n=19). Both groups were compared to age-match controls (n=20) who had no neurological impairment/disease, and no history of head trauma, either by playing contact sports, or trauma from accidents. All data was completed during one visit to the laboratory and cognitive and TMS testing was randomised to reduce any potential serial order effects.

### Symptom self-report

All participants completed a questionnaire regarding their concussion injury history (Pearce et al., 2014), and a self-assessment regarding fatigue and related concerns affecting their daily activities (Johansson et al., 2009). The self-assessment required participants to respond to 15 questions covering a range of concerns including fatigue (general and mental), perception of thinking speed and mental recovery, emotional, irritability and sensitivity changes, and sleep variability, using a Likert rating scale from 0 to 3, in 0.5 increments. Higher scores reflect greater severity for each symptom-related question. The questionnaire has been previously validated by Johansson and colleagues (2009, 2010, 2014).

### Cognitive assessment

Participants completed a computerised brief battery (Cogstate, Melbourne, Australia) that comprised of a subset of tasks from the full Cogstate battery taking about 8-10 minutes in total (Maruff et al., 2013). Prior to data collection participants were given a five-minute interactive demonstration and familiarisation. Once participants had demonstrated they were aware of the assessment protocol, data collection began.

Participants completed two separate reaction time tests; a simple reaction time ‘detection test’ where the individual was instructed to respond as quickly as possible by pressing a keyboard key as soon as the card was revealed (‘turned up’), and a choice reaction time ‘identification test’ where the participant pressed one of two keys; one representing the ‘yes’ button if the card was revealed red in colour, or another key representing the ‘no’ button if the card was black in colour. For both the detection and identification assessments, if a key was pressed before the card was revealed, this would be recorded as an error, contributing to the accuracy metric. The test was completed when 25 correct responses were recorded or the maximum time (three min) had elapsed (Maruff et al., 2009).

Tests for response times included the One-Back and Visual Learning tasks. The one-back task required the participant to respond to the question “is this card the same as the previous card?” Participants were instructed to press a particular key for a ‘yes’ or an alternative key for a ‘no’ response as soon as possible. Cards (n=42) were shown, and the correct response was 50% each of the trials presented. The test was completed when all 42 trials were completed or the maximum allowed time of three minutes had passed (Maruff et al., 2009). The visual learning tasked required the participant to view the card presented in the middle of the screen and respond to the question “have you seen this card before?” Similar to the one-back task, participants were instructed to press a particular key for a ‘yes’ or an alternative key for a ‘no’ response. Participants were required to learn a series of six cards repeated throughout the task, intermixed with eight non-repeating ‘distracter’ cards in series of 14 cards. Three 14-card series were presented, and this task continued until the participant had made 42 complete responses or the maximum time allowed (3 min) had elapsed. The primary outcome measure for this task was the number of correct responses (i.e., true-positive and true-negative) expressed as a proportion of the total trials (Maruff et al., 2009).

### Somatosensory assessment

As described in previously published studies (Pearce et al., 2019; M. Tommerdahl et al., 2016; Zhang et al., 2011) somatosensory assessment was undertaken by utilising a portable vibrotactile stimulation device (Brain Gauge, Cortical Metrics, USA). Physically similar to a standard computer mouse, the device contains two cylindrical probes (5 mm diameter) positioned at the top and front of the device. These probes, driven by the computer via a USB cable, provided a light vibration stimulus, at frequencies between 25–50 Hz that is sensed by the participant’s index and middle digits of their non-dominant hand.

Participants completed the battery involving four discrete tasks, one reaction time and three discriminative tasks (amplitude, duration and temporal order judgement), whereby the participant used their non-dominant hand to detect the stimulus, and their dominant hand to respond via a computer mouse. Testing time took approximately 15 minutes. For the discrimination tasks in the battery, a simple tracking procedure that utilized a two-alternative forced choice paradigm was used to determine an individual’s difference distinguished threshold for stimulus (M. Tommerdahl et al., 2016).

Familiarization was performed before each test for participant orientation, requiring correct responses on three consecutive trials before progressing the test where data would be acquired. Participants were verbally instructed to respond as quickly as possible, and during testing no feedback or knowledge of the results were provided.

### Corticospinal excitability

Employing previously published methods in similar cohorts (Pearce et al., 2014; Pearce et al., 2021; Pearce et al., 2018), corticospinal excitability was quantified via single-pulse TMS, delivered over the contralateral primary motor cortex. Surface electromyography (sEMG) measured motor evoked potentials (MEPs) recording 500 ms sweeps (100 ms pre-trigger, 400 ms post-trigger; PowerLab 4/35, ADInstruments, Australia). Electromyography, adhering to the Non-Invasive Assessment of Muscles (SENIAM) guidelines for sEMG (Hermens et al., 1999), was recorded using bipolar Ag/AgCl electrodes, with an intra-electrode distance of 2 cm positioned over the first dorsal interosseous (FDI) muscle of the participant’s dominant hand, and the ground electrode placed over metacarpophalangeal joint of the third digit.

Single pulse TMS was delivered using a MagStim 200^*2*^ stimulator (Magstim, UK) and a figure-of-eight coil (Magstim, UK). Reliability of coil placement was maintained by participants wearing a snugly fitted cap (EasyCap, Germany), positioned with reference to the nasion-inion and interaural lines. The cap was marked with sites at 1 × 1 cm spacing in a latitude-longitude matrix to provide reliable coil position throughout the testing protocol (Pearce et al., 2000).

Following identification of the ‘optimal site’, defined as the site with the largest observed MEP (Pearce et al., 2000), active motor threshold (aMT) was determined via a low-level voluntary static contraction of the FDI muscle at 10% of Maximal Voluntary Contraction (MVC). The aMT was identified by delivering TMS stimuli (5% of stimulator output steps, and in 1% steps closer to threshold) at intensities from a level below the participant’s threshold until an observable MEP of at 200 *µ*V and associated cSP could be measured in at least five of ten stimuli (Pearce et al., 2013; Wilson et al., 1995). Once aMT was established, 20 stimuli (four sets of five pulses per set) were delivered in random intervals (between 6–10 s) at intensities to evoke a MEP of 1 mV. A break of 30 s was provided between sets to reduce any possibility of muscular fatigue (Kidgell & Pearce, 2010).

### Data and statistical analyses

Self-report symptom score were totalled from the responses of the 15 questions, giving a maximum of 44 points (Johansson et al., 2009). Outcome measures from Cogstate included percentage of correct responses and mean reaction time for the detection test, and mean response time for the identification test, One-Back and Visual Learning tasks (Maruff et al., 2009). For the somatosensory testing, apart from the mean reaction time for the detection of the sensory stimulus, the discrimination assessments measured response time and calculated score for following presentation of the stimulus (King et al., 2018; A. Tommerdahl et al., 2019). Single pulse MEP latency was calculated as the time between stimulation of the motor cortex to the onset of the MEP (Brasil-Neto et al., 1992). MEP amplitudes were measured from the peak-to-trough difference of the waveform. Duration of the cSP was calculated from the onset (deflection) of the MEP waveform to the return of uninterrupted EMG (Wilson et al., 1993). With the most influencing confounding factor on cSP duration being the preceding MEP (Škarabot et al., 2019), we employed MEP:cSP ratio to compare between groups and reduce between-participant variability (Orth & Rothwell, 2004). We have previously published MEP:cSP ratios in a cohort with persistent post-concussion symptoms (Pearce et al., 2020) and more recently in larger project on retired contact sport athletes (Pearce et al., 2021).

All statistical analyses were conducted using Jamovi software (www.jamovi.org, Version 1.0.8). Data were tested for normality using Shapiro-Wilks (S-W) tests showing data to be skewed (all variables *p*<0.05). Data were analysed using Kruskal-Wallis tests with Dwass, Steel, Critchlow-Fligner post-hoc comparisons, except for comparison for competitive career, the number of concussions, and time since last concussion between ‘self-concern’ and ‘no-concern’ groups which was analysed using a Mann-Whitney test. Effect sizes are presented as rank-biserial correlation (*r*_*rb*_) for 2-group or partial eta squared (*n*^*2*^*p*) for 3-group comparisons. The number of previous concussions and the fatigue and related symptom scores to cortical metrics and TMS variables were correlated using Kendall’s Tau B. Data in Tables and Figures are presented as mean (± SD) and statistical significance as set as alpha <0.05.

## Results

There were no difference in participant age (*H*(2)=0.61, *p*=0.74, *n*^*2*^*p*=0.01), and education (*H*(2)=1.89, *p*=0.11, *n*^*2*^*p*=0.23) between all groups. Between retired athlete groups there was no difference in career length (*U*=242, *p*=0.33), the number of concussions (*U*=263, *p*=0.33), or time since last reported concussion (*U*=233, *p*=0.34; **Table 1**).

**Table 2** presents all items of the fatigue and related symptom questionnaire. There were significant differences observed between groups for total score (*H*(2)=63.27, *p*<0.001, *n*^*2*^*p*=0.85). Post hoc comparisons showed the group reporting ongoing concerns with their mental or cognitive health (‘self-concern’) had significantly higher total scores than both control participants (*W*=8.72, *p*<0.001), and those ex-players with no ongoing concerns (‘no concern’; (*W*=8.57, *p*<0.001). This pattern was seen in almost every item within the survey except for sensitivity to light and noise, where post hoc differences were observed between ongoing concerns and no concerns groups, and ongoing concerns and control groups (*p*<0.001). For decreased sleep, differences were found only between the ongoing concern and control groups (*p*<0.001). While the no concern group rated higher on the decreased sleep compared to controls, this was not statistically significant (*p*=0.103). There was no difference in increased sleep between groups (*H*(2)=4.51, *p*=0.105). The total fatigue and related symptom score was not correlated to age (Kendall’s Tau *B*=0.11, *p*=0.26), nor to the number of concussions (Kendall’s Tau *B*=0.005, *p*=0.96), nor the time since last concussion (Kendall’s Tau *B*=0.12, *p*=0.28).

**Table 2.** Fatigue and related symptoms scores (mean ± SD)

|  | General<br>fatigue | Lack<br>initiative | Mental<br>fatigue | Mental<br>recovery | Concentration<br>difficulties | Memory<br>problems | Slowness<br>thinking | Sensitive<br>to stress | Emotional<br>instability | Irritability | Sensitivity<br>to light | Sensitivity<br>to noise | Decreased<br>sleep | Increased<br>sleep | Total<br>score |
| --- | --- | --- | --- | --- | --- | --- | --- | --- | --- | --- | --- | --- | --- | --- | --- |
| **Self-**<br>**concern**<br>**(n=36)** | 1.81<br>$\pm 0.66^{1,4}$ | 1.65<br>$\pm 0.45^{1,4}$ | 2.11<br>$\pm 0.41^{1,3}$ | 1.41<br>$\pm 0.82^{1,4}$ | 1.92<br>$\pm 0.52^{1,3}$ | 1.93<br>$\pm 0.55^{1,3}$ | 1.89<br>$\pm 0.61^{1,3}$ | 2.27<br>$\pm 0.80^{1,3}$ | 1.77<br>$\pm 0.92^{1,3}$ | 2.07<br>$\pm 0.75^{1,4}$ | 1.41<br>$\pm 0.81^{1,4}$ | 1.61<br>$\pm 0.78^{1,3}$ | 1.81<br>$\pm 0.90^1$ | 0.64<br>$\pm 1.03$ | **25.32**<br>**$\pm 4.76^{1,3}$** |
| **No**<br>**concern**<br>**(n=20)** | 1.26<br>$\pm 0.54^1$ | 1.03<br>$\pm 0.59^2$ | 1.39<br>$\pm 0.66^1$ | 0.84<br>$\pm 0.60^1$ | 1.32<br>$\pm 0.51^2$ | 1.21<br>$\pm 0.56^2$ | 1.05<br>$\pm 0.52^2$ | 1.53<br>$\pm 0.61^1$ | 0.87<br>$\pm 0.50^2$ | 1.37<br>$\pm 0.81^2$ | 0.71<br>$\pm 0.67$ | 0.89<br>$\pm 0.46$ | 1.21<br>$\pm 0.98$ | 0.58<br>$\pm 0.63$ | **15.24**<br>**$\pm 2.33^1$** |
| **Control**<br>**(n=19)** | 0.56<br>$\pm 0.57$ | 0.36<br>$\pm 0.51$ | 0.33<br>$\pm 0.66$ | 0.19<br>$\pm 0.35$ | 0.61<br>$\pm 0.74$ | 0.67<br>$\pm 0.71$ | 0.61<br>$\pm 0.76$ | 0.67<br>$\pm 0.86$ | 0.47<br>$\pm 0.67$ | 0.75<br>$\pm 1.05$ | 0.50<br>$\pm 0.62$ | 0.47<br>$\pm 0.63$ | 0.67<br>$\pm 0.73$ | 0.19<br>$\pm 0.52$ | **4.82**<br>**$\pm 2.74$** |

<sup>1</sup> Significance vs control group ( $<0.01$ ); <sup>2</sup> Significance vs control group ( $<0.05$ ); <sup>3</sup> Significance vs no concern group ( $<0.01$ ); <sup>4</sup> Significance vs no concern group ( $<0.05$ )

Cognitive assessment revealed no differences in accuracy between groups in each of the four the Cogstate tests performed (**Figure 1a**). However, the time taken to respond to the questions in three of four of these tests was significantly longer in the group with ongoing self concern (**Figure 1b**). Reaction times for the visual detection and attention, tasks showed significant differences (*H*(2)=10.61, *p*=0.005, *n*^*2*^*p*=0.14) with post hocs revealing a significantly greater time in the group with self concern than both no concern (*W*=3.88, p=0.017) and control groups (*W*=3.88, p=0.017), despite near identical accuracy scores. Response times in the visual learning task was significantly longer between groups (*H*(2)=11.32, *p*=0.003, *n*^*2*^*p*=0.15) and post hoc comparison showing a significant difference in response times with ongoing concerns relative to ex-players with no ongoing concern (*W*=3.60, p=0.029) and controls (*W*=3.94, p=0.015; **Figure 1b**). No differences were detected between groups in the response time of the working memory task (*H*(2)=4.61, *p*=0.1, *n*^*2*^*p*=0.06). The time taken to respond in all cognitive tasks was positively correlated to fatigue score, however the task result (i.e. the accuracy of response) was not (**Table 3**).

**Figure 1.**
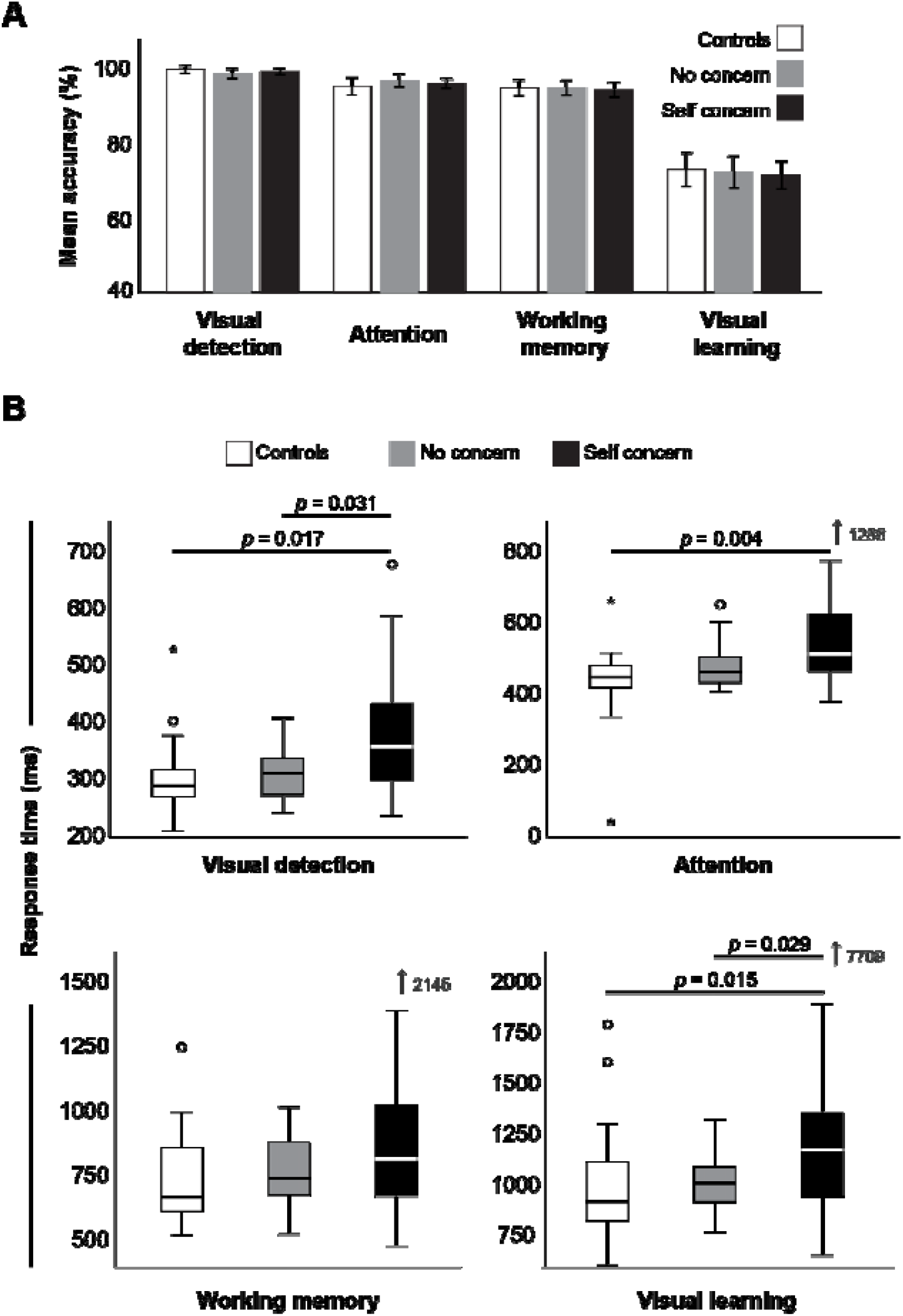
CogState accuracy (a) and median response times between groups.

**Table 3.** Correlation^a^ between self-reported fatigue score and objective measures

|  | **Test**<br>**score** | ***p*** | **Response**<br>**time** | ***p*** |
| --- | --- | --- | --- | --- |
| **Visual detection** | -0.042 | ns | 0.278 | <□0.001 |
| **Attention** | 0.031 | ns | 0.277 | <□0.001 |
| **Visual learning** | -0.017 | ns | 0.244 | 0.002 |
| **Working memory** | -0.072 | ns | 0.172 | 0.031 |
| **Sequential Amplitude** | -0.029 | ns | 0.177 | 0.027 |
| **Simultaneous Amplitude** | 0.085 | ns | 0.204 | 0.01 |
| **Duration Discrimination** | 0.022 | ns | 0.211 | 0.008 |

<sup>a</sup> Kendall's Tau B correlation coefficient

Like cognitive testing, somatosensory testing using Cortical Metrics showed no difference in the mean score between groups (**Figure 2a**), but again, response times were consistently longer in the self concern group (**Figure 2b**). Specifically, there were significantly delayed reaction times in sensory detection, and response times for sequential amplitude, simultaneous amplitude, and duration discrimination, relative to the control group. Again response times, not overall scores, were significantly positively correlated with fatigue scores (**Table 3**).

**Figure 2.**
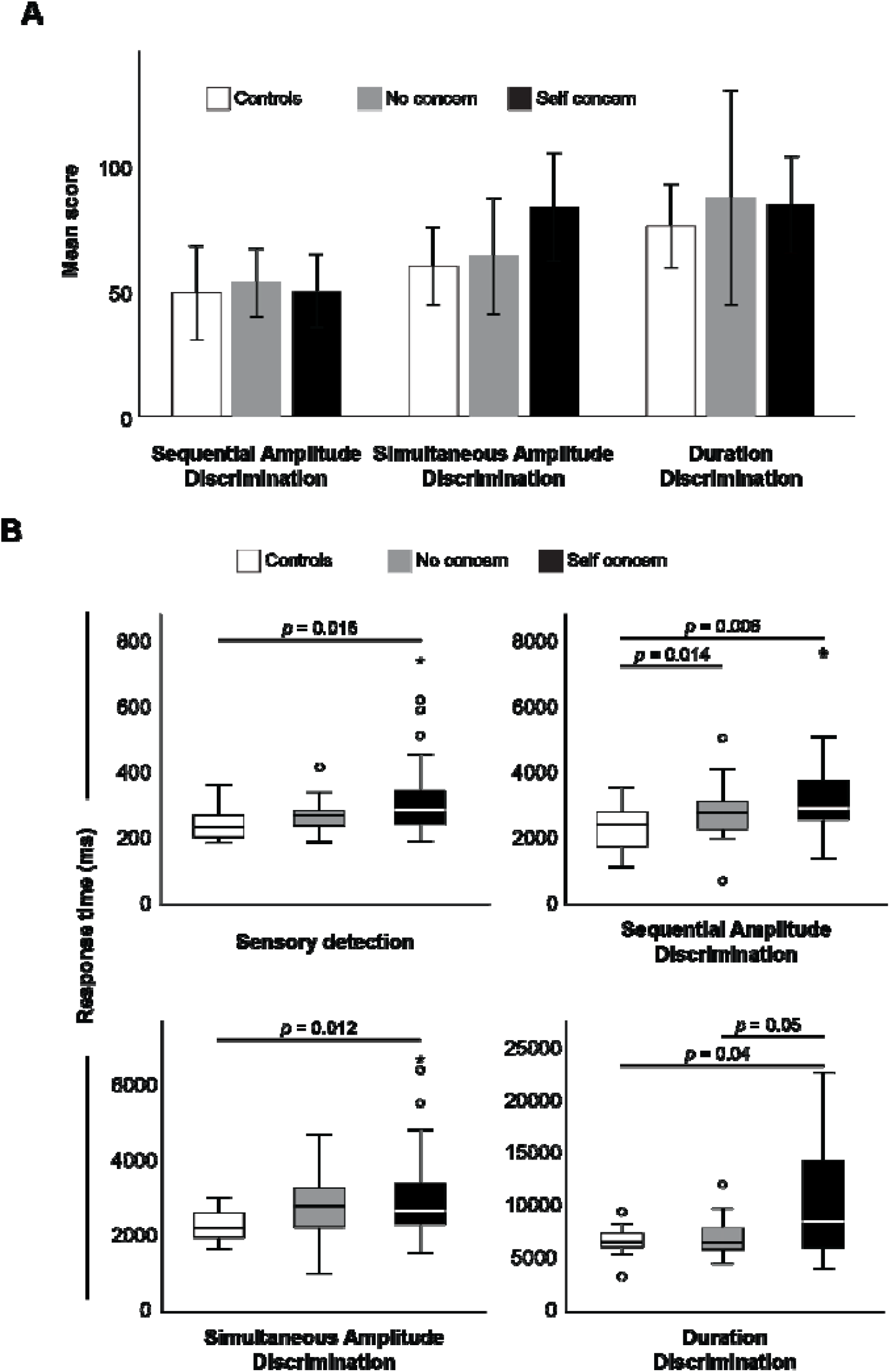
Mean score (a) and response times (b) between groups.

Differences among groups were also found during transcranial magnetic stimulation (TMS). While the median MEP:cSP ratio (**Figure 3a**) was increased in ex-players both with self concern and no concern, compared to control (37.8 and 48.0 v 23.6, respectively), this did not reach statistical significance. However, MEP latency was significantly prolonged in the players with self concern relative to control (*H*(2)=9.73, *p*=0.008, *n*^*2*^*p*=0.13; **Figure 3b**), suggesting the presence of damage to motor pathways in this group that cannot be discerned from MEP amplitudes alone. TMS latency was, like the response times of cognitive and somatosensory tests, significantly positively correlated with fatigue score (Kendall’s Tau B = 0.182, *p* = 0.024).

**Figure 3.**
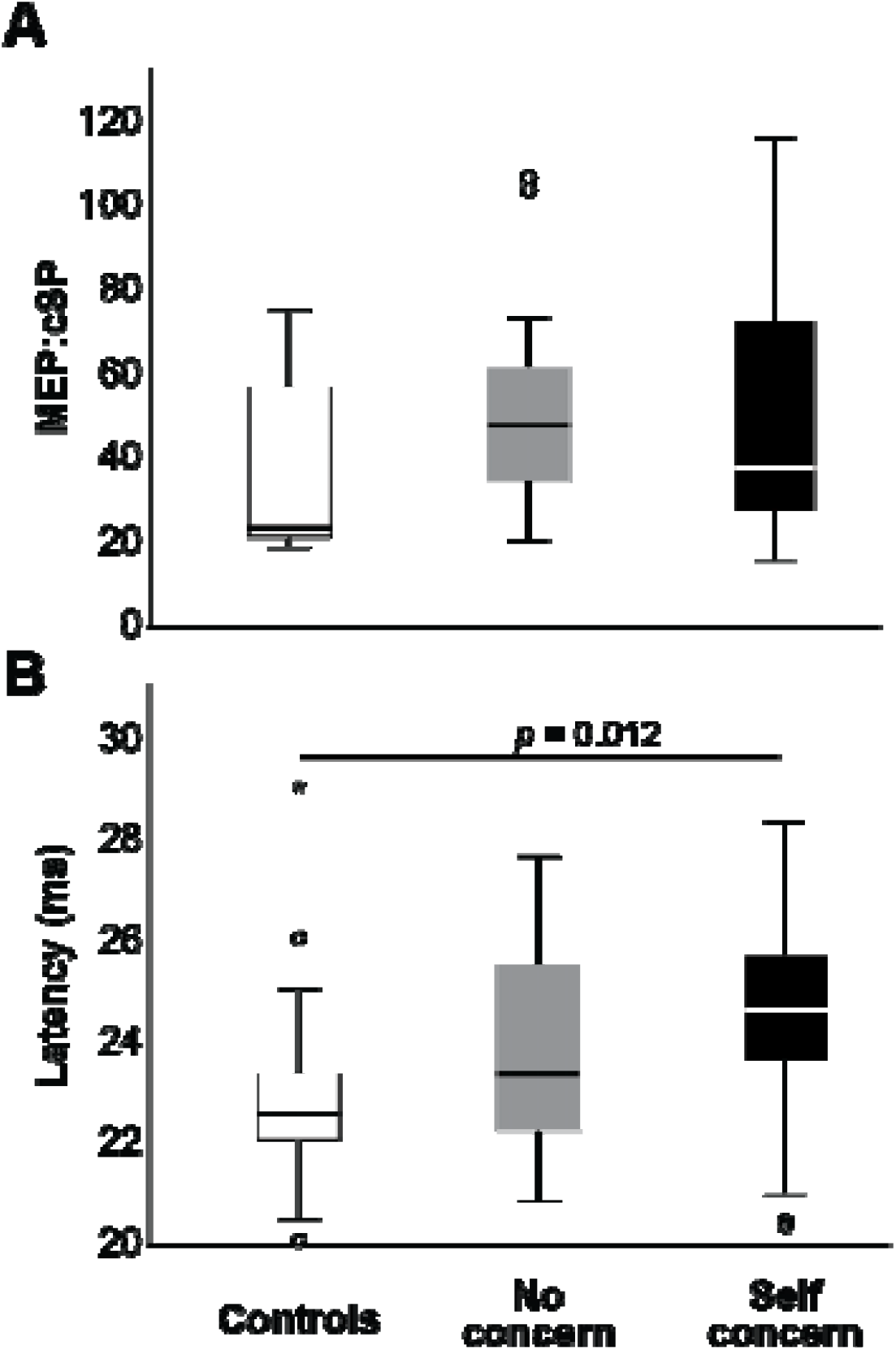
Mean TMS MEP:cSP ratio (a) and MEP latency (b) between groups.

## Discussion

Our study has found that retired contact sport athletes with self-concerns were significantly slower in both reaction time and response time, compared to retired athletes with no concerns, and age-matched controls. Moreover, reaction and response times correlated with self-reported fatigue and related symptom scale total score, and corticospinal latency. While groups did not differ in outcome performance (i.e. accuracy), the difference in reaction and response times suggest a lack of efficiency at processing ability (A. Tommerdahl et al., 2019) which appeared unrelated to sleep concerns (i.e. the two retired athlete groups did not differ in either decreased in increased sleep concerns). We consider the slowing of responses in lieu of accuracy scores an important finding as the majority of studies investigating cognitive health outcomes in these cohorts report performance outcomes, with only a minority presenting abilities via psychomotor reaction times (Cunningham et al., 2020). Moreover, the novel finding of impaired response times suggests that cognitive impairment of retired athletes with a history of head trauma should include response times in future studies.

We have previously employed the fatigue and related symptom survey to characterise and quantify our cohorts, particularly those who express ongoing self-reported symptoms compared to those who report no ongoing symptoms (Pearce et al., 2020; Pearce et al., 2021). We found significant differences between the groups we studied, with the self-concern group having the highest scores. However we also found that players with no-concern scored on average above the 10.5 clinical cut off score for “normal” as suggested by Johansson and Rönnbäck (2014). This may imply that there is an underlying clinical issue for the individual, although a serious problem whereby activities of daily living are significantly affected, is not always the case. For this study, our sample was derived from those who volunteered for testing who explicitly expressed they had no ongoing concerns, and we used the total score to characterise between groups.

Slowed response times in the acute period (one to two weeks) following a concussion injury has been previously reported (A. Tommerdahl et al., 2019), but to the best of our knowledge this is the first study to report slowed response times in a long-term cohort with a history of repeated head trauma. While the visual learning task was not statistically significant, the response times reflected the same pattern as the other reaction time and response time tasks: the control group showed the fastest while the self-concern group showed the slowest. Coupled with the TMS data demonstrating altered corticospinal latency, the data suggests that those with a history of repeated neurological insults have some effects on processing ability, with those reporting greater severity of symptoms reflected in worse response times and significantly reduced corticospinal excitability ratio.

Our previous studies primarily focussed on neurophysiological alterations in both persistent post concussion symptoms (Pearce et al., 2020; Pearce et al., 2019) and chronic outcomes of repeated head trauma (Pearce et al., 2014; Pearce et al., 2021; Pearce et al., 2018). In contrast, this study was aimed to quantify response times, while TMS was used to provide a potential physiological mechanism to explain differences between groups (De Boeck & Jeon, 2019). While we acknowledge that TMS is an indirect measure of corticospinal excitability and latency is a raw measure of conduction speed, the correlations between significantly prolonged corticospinal latency and cognitive response times in the ‘self-concern’ group was surprising. However, this is not the first time that slowed TMS latency has been reported. Livingston et al reported a slowing of TMS latency in the acute phase following a concussion (Livingston et al., 2012; Livingston et al., 2010), while Stokes et al. (2020) recently reported increased TMS latencies in young athletes (18-22 years) who had reported a history of concussions (>1 year). Our findings, in a group of older retired athletes, may reflect alterations in white matter in the pyramidal pathways (Stokes et al., 2020), where MEP latencies have been shown to increase with demyelination associated with neurogenerative disease (Britton et al., 1991; Schmierer et al., 2002); it has also been postulated that slowed conduction time may be due to neurochemical changes associated with a history of physical brain trauma (Lin et al., 2015). While further research, particularly studies where co-registration of TMS and neuroimaging can be performed, is required, employing response times in computerised cognitive-motor and sensorimotor testing, along with low-cost physiological techniques such as TMS may provide a more accurate picture of long-term cognitive health concerns in those with a history of repeated head trauma.

It is outside of the scope of the study to speculate on why some of the retired playing cohort were more affected than others in their self-report. However, the aim of the study was to address the concerns regarding potentially biased sampling that has previously been suggested (Carman et al., 2015). Similar to our more recent studies (Pearce et al., 2021) we specifically aimed to recruit retired athletes with a history of head trauma both with and without ongoing self-reported concerns. In line with our previous work, we found that the group with no reported symptoms fared significantly better than the group with self-reported concerns but did show small-to-moderate effects compared to the age-matched control group. Collectively these data shows that repeated head trauma may affect cortical processing, however there may be a ‘threshold’ before this becomes a clinical concern. Further research is required to ascertain what this threshold may be from a physiological perspective.

There are several limitations to consider in this study. Firstly, we have relied on self-report for participants’ concussion history. To assist with recollection we used the criteria of missing playing the following week (AFL Medical Officers Association, 2011), however, this may still underestimate the number of concussions players experienced. Moreover, while we report similar career lengths between the two retired playing groups, we are not able to consider the exposure of repetitive sub-concussive trauma experienced, that did not result in concussion signs or symptoms, that may contribute to the neural degradation suggested by increased TMS latency data. Secondly, similar to previous work (De Beaumont et al., 2009; Pearce et al., 2014; Pearce et al., 2021; Pearce et al., 2018), this study used a retrospective cross-sectional design. While we were not able to obtain data of the players’ pre-morbid functioning, we aimed to address this by having a three-group design incorporating an ‘active-control’ group of retire players with a similar history of reported concussions, but no ongoing concerns. Future studies would benefit from prospective designs with players being tested prior to starting their careers, but in light of current cohorts, future studies should consider repeated measures to quantify time-related progressive changes between groups of currently retired athletes.

In conclusion, this study is the first to present slowed response times in a cohort of older, retired contact sport athletes with ongoing concerns regarding their head trauma history. While outcome results did not differ between groups, the finding of poorer response time performance, suggests less cognitive processing efficiency and neural conduction integrity, and may underpin the concerns, expressed by some retired players, with regards to struggling with activities of daily living. With computerised testing that collects response time data, our data suggest that analyses of cognitive health will be more informative with the inclusion of cognitive-motor and/or sensorimotor response times.

## Supporting information

CONSORT

## Data Availability

All data produced in the present study are available upon reasonable request to the authors approved by the requester's research institution.

## Acknowledgement/Declarations of Interest

This study did not receive any specific grant from funding agencies in the public, commercial, or not-for-profit sectors. AJP currently receives partial research salary funding from Erasmus+ strategic partnerships program (2019-1-IE01-KA202-051555). AJP has previously received partial research funding from the Sports Health Check Charity (Australia), Australian Football League, Impact Technologies Inc., and Samsung Corporation, and is remunerated for expert advice to medico-legal practices. The development and manufacture of the Cortical Metrics device used in this study has received partial funding from the Office of Naval Research (USA). MT is a director of Cortical Metrics LLC who has a license from the University of North Carolina to distribute the Brain Gauge device used in this study. No other author has any declaration of interest.

